# Sex-differences in reporting of statin-associated diabetes mellitus to the US Food and Drug Administration

**DOI:** 10.1101/2024.05.01.24306727

**Authors:** David P Kao, James Martin, Christina Aquilante, Elise L Shalowitz, Katarina Leyba, Elizabeth Kudron, Jane EB Reusch, Judith G. Regensteiner

**Affiliations:** University of Colorado Ludeman Family Center for Women’s Health Research, University of Colorado School of Medicine, Aurora, CO; Department of Medicine, University of Colorado School of Medicine, Aurora, CO; Department of Biomedical Informatics, University of Colorado School of Medicine, Aurora, CO; Colorado Center for Personalized Medicine, Aurora, CO; Department of Pharmaceutical Sciences, University of Colorado Skaggs School of Pharmacy & Pharmaceutical Sciences, Aurora, CO; Department of Pediatrics, University of Colorado School of Medicine, Aurora, CO; Rocky Mountain Regional VA Medical Center, Aurora, CO

## Abstract

**Objective:** Determine whether statin-associated DM is reported more frequently in women than men in post-marketing adverse drug event (ADE) surveillance.

**Design:** Retrospective pharmacovigilance analysis

**Data source:** Publicly available FDA Adverse Event Reporting System (FAERS) from January 1997 through December 2023.

**Setting:** Real-world spontaneously reported ADEs in the United States.

**Participants:** Community patients reporting statin ADEs during the study period.

**Interventions/exposures:** Adverse drug event reports that included at least one statin.

**Main outcome measures:** Proportional reporting ratio to identify increased rates of statin-associated DM events in women and men compared with all other medications, and reporting odds ratio to compare reporting rates in women vs. men.

**Results:** A total of 18,294,814 ADEs were reported during the study period. Among statin-associated ADEs, 14,897/519,209 (2.9%) reports mentioned DM in women compared with 7,412/489,453 (1.5%) in men, which were both significantly higher than background (0.6%). Statins were primary-or secondary-suspected cause of the ADE significantly more often in women than men (59.8 vs. 28.7%), and reporting rates were disproportionately higher in women than in men for all statins. (reporting odds ratio 1.9 [95% CI 1.9-2.0]). The largest difference in reporting of statin-associated DM between women and women was observed with atorvastatin.

**Conclusions:** Analysis post-marketing spontaneous ADE reports demonstrated a higher reporting rate of DM-associated with statin use compared to other medications with a significantly higher reporting rate in women compared to men. Future studies should consider mechanisms of statin-associated DM moderated by sex.

## INTRODUCTION

High cholesterol is a common cardiovascular risk factor in the US and internationally. ^1^ Statins have been among the most prescribed class of drugs with an estimated 92 million patients on statin therapy in 2019 in the US alone. ^2^ Statins have been shown to have generally good safety with comparable efficacy between men and women in terms of lipid-lowering effect and reduced onset of atherosclerotic cardiovascular disease. Despite this, a number of sex differences in the utilization of statins have been observed ranging from differences in prescribing rates and statin dose to safety profile and long-term adverse effects. ^3–7^ Optimization of lipid-lowering therapy must therefore consider efficacy and safety in choosing agent, dose, and perhaps even class of medication in the context of patient’s sex.

Sex differences in statin-induced diabetes mellitus (DM) were first suggested by Culver, et al. in the Women’s Health Initiative, wherein the odds ratio (OR) of developing DM across all statins was ∼ 1.7. ^8^ In comparison, randomized clinical trials involving statins comprised predominately of men showed lower ORs ranging from 0.95 – 1.14.^9^ The mechanisms of statin-induced DM are unclear but may include decreased insulin production by pancreatic β-cells, decreased production of ubiquinone, decreased GLUT4 adipocytes, or inhibition of glucose-mediated insulin release in cells.^10^

Awareness of DM as a potential consequence of statin use is growing and is now included in patient-facing educational resources from agencies like the Centers for Disease Control.^11^ Despite growing acceptance of DM as an adverse side effect of statins, there is relatively little data directly comparing risk between men and women. Herein we summarize a pharmacovigilance analysis that identifies signals of disproportionate reporting (SDR) of sex differences of statin-associated DM in the Food and Drug Administration’s Adverse Event Reporting System (FAERS). We hypothesized that statin-associated DM is reported disproportionately more in women than in men in post-marketing adverse drug event (ADE) surveillance representing an important sex difference in the safety profile of this commonly used class of medications.

## METHODS

### IRB approval

All data are deidentified and available for unrestricted public download. Our analysis therefore not require ethics approval.

### Patient and Public Involvement

Patients and the public were involved in this study through voluntary, spontaneous submission of ADEs to the FDA, either directly or via the drug manufacturer. Patients and providers can currently report ADEs via the MedWatch website (https://www.accessdata.fda.gov/scripts/medwatch/). There was no direct patient/public content as part of this study, and all data were fully deidentified prior to download.

### Data sources

We obtained publicly available data from FAERS (https://open.fda.gov/data/faers/). Sex is not available in the FAERS data prior to 1997. We therefore analyzed ADE reports received from January 1997 through December 2023. ADEs in FAERS have two separate identifiers. The *PRIMARYID* identifier reflects a specific report submitted, e.g. through MedWatch. The *CASEID* identifier denotes a unique clinical event, which may be described by multiple individual reports from different sources. To avoid redundant counting, we generated results using unique case identifiers (*CASEID*).

### Data preprocessing

FAERS ADE reports include a list of all drugs present in each ADE, which may include trade names, combination drugs, and obsolete or foreign names. Therefore, all FAERS cases were matched with distinct pharmacologic components using a multi-tiered algorithm of whole and partial string matching as well as limited manual matching by the authors. Drugs@FDA was used as the reference for drug brand names and active ingredients as described in FAERS documentation. ^12^ Using this approach, more than 95% of all FAERS reports were matched to at least one active ingredient from Drugs@FDA. Unmatched names (e.g. “HC”, ”cranberry”, ”BLU-U Blue light photodynamic therapy illuminator”) were excluded from this analysis. Access to this transformed dataset can be provided via comma-separated value files or access to our Google BigQuery repository on reasonable request.

FAERS uses Medical Dictionary for Regulatory Activities (MedDRA, version 25.1) Preferred Terms (PT) to classify ADEs. The specific PTs used to identify DM in this analysis were “diabetes mellitus”, “diabetes mellitus inadequate control”, “diabetes with hyperosmolarity”, “diabetes complicating pregnancy”, “gestational diabetes”, “increased insulin requirement”, “insulin resistant diabetes”, “pancreatogenous diabetes”, “insulin-requiring type 2 diabetes mellitus”, “type 1 diabetes mellitus”, “type 2 diabetes mellitus.”

### Statistical analysis

The primary analysis used the aggregate of all approved statins combined (atorvastatin, fluvastatin, lovastatin, pitavastatin, pravastatin, rosuvastatin, simvastatin). We did not include cases involving cerivastatin. Analyses were repeated for each of the four most reported statins (atorvastatin, pravastatin, rosuvastatin, simvastatin, **Figure 1**). The primary outcome was the presence of a significant difference between men and women in the spontaneous reporting rate of DM associated with all statins combined. Secondary outcomes were sex differences in spontaneous reporting of DM associated with each individual statin.

**Figure 1.**
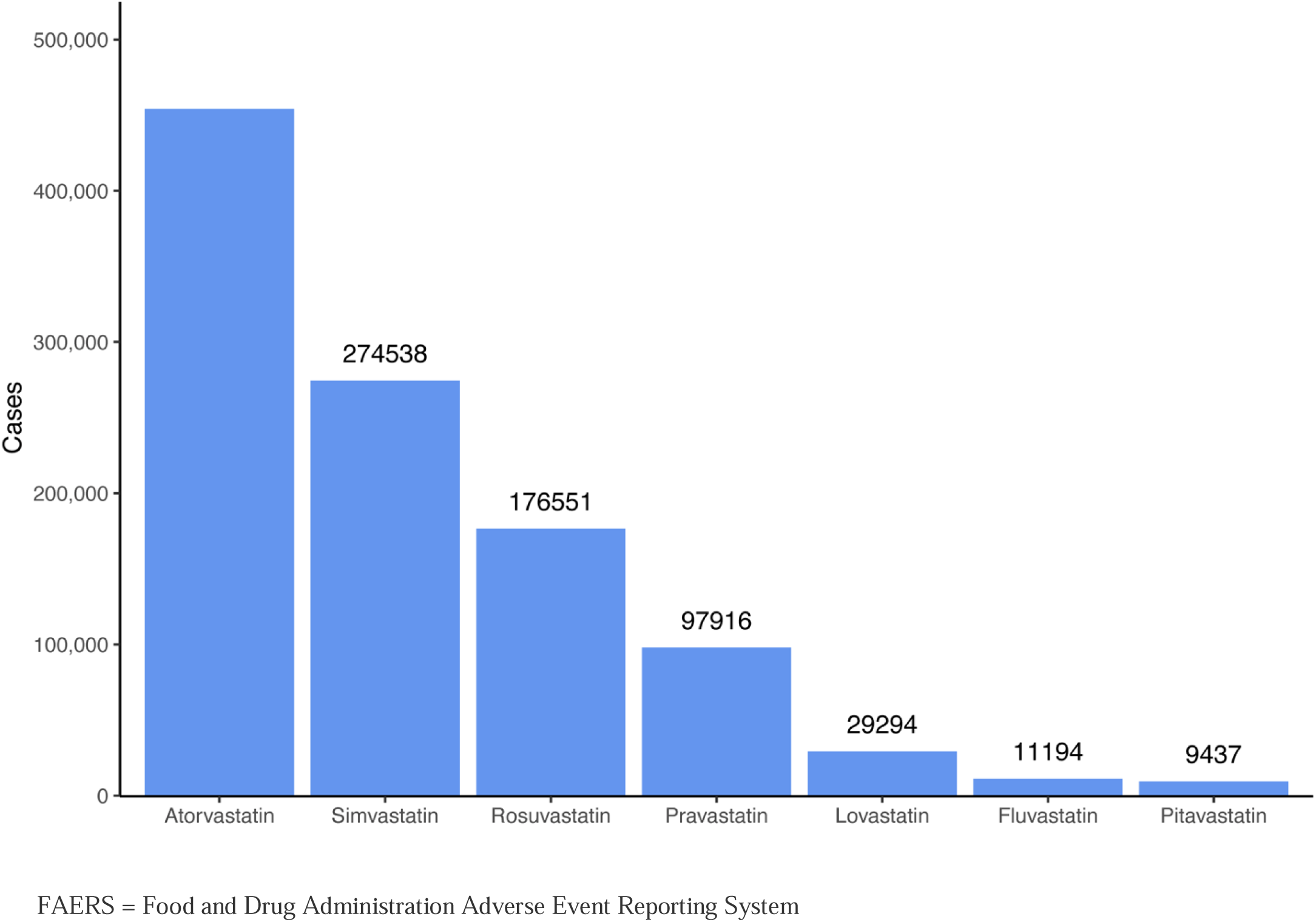
Total unique statin cases in FAERS, 1/1/1997-12/31/2023.

Pharmacovigilance analyses of spontaneously reported ADE data like those from FAERS generally involve identifying SDRs, wherein certain ADEs are reported more frequently with the drug(s) of interest compared with the reporting frequency of the same ADE in all other reports combined. When studying large numbers of events, the Proportional Reporting Ratio (PRR) has been used and recommended by drug safety monitoring agencies such as the FDA, Eudravigilance, and the UK’s Yellow Card Scheme.^13^ We therefore used the PRR as the primary indicator of an SDR for DM associated with statins compared to all other drugs. The PRR was considered significant if the PRR point estimate was > 2, χ^2^ ≥ 4, and there were ≥ 3 reports of DM associated with statins.^14^ The PRR is calculated as follows:

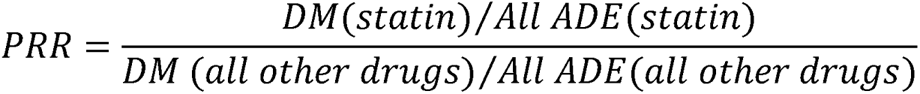

We then tested for differences between reporting in men and women using the Reporting Odds Ratio (ROR), which has used in prior analyses to comparing SDR in subgroups.^15^ A difference in statin-associated DM reporting was considered significant if the lower bound of the ROR 95% confidence interval (CI) was > 1 and there were ≥ 3 reports of DM associated with statins.^13^ The ROR was calculated as follows:

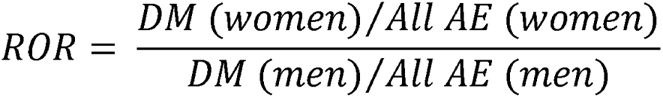

Categorical and continuous variables were otherwise compared using χ^2^ analysis, and analysis of variance respectively. A p-value < 0.05 was considered significant for both. All analyses were performed using RStudio (version 2023.12.0, Posit Software, Boston, MA) and the R statistical package (version 4.3.2, R Foundation for Statistical Computing, Vienna, Austria). SDR analyses were performed using the *mdsstat* package (version 0.3.2, ASM Inc., Temecula, CA). All FAERS, Drugs@FDA, and MedDRA data are hosted for analysis in Google BigQuery (cloud.google.com, Mountain View, CA). Analysis code and access to our FAERS BigQuery repository are available on reasonable request.

## RESULTS

### All statins

There were 18,294,814 unique ADE reports during the study period, of which 121,372 (0.7%) reported DM as defined above and 1,085,700 cases mentioned at least one of the 7 statins listed above. The distribution of statin reports during the study period is summarized in **Figure 1**. Sex was reported in 1,008,665 (92.9%) statin cases with 519,209 (51%) involved women. Sex was not reported in 77,035 (7.1%) cases.

Characteristics of all reported cases of statin-associated DM are found in **Table 1**. There were 2-fold as many reports of statin-associated DM in women vs. men (14,794 vs. 7,412, respectively) compared with only 6% more statin-associated reports of non-DM ADEs (519,209 vs. 489,456, respectively). Statins were more frequently primary or secondary suspect in ADEs reporting DM in women than men (71 vs. 30%, p<0.001), although reported rates of death (2% vs. 9%) and hospitalization (15% vs. 38%) were significantly higher in men (p<0.001 for both).

**Table 1.** Characteristics of statin-associated DM for all statins, 1997-2023.

|  | **Female**<br>**(N=14,897)** | **Male**<br>**(N=7,412)** | **p-value** |
| --- | --- | --- | --- |
| Age, years | 58.6±11.6 | 59.9±13.0 | <0.001 |
| **Suspected role of drug\*** |  |  |  |
| Primary suspect | 7931 (53.2%) | 1244 (16.8%) | <0.001 |
| Secondary suspect | 976 (6.6%) | 882 (11.9%) |  |
| Interacting | 17 (0.1%) | 26 (0.4%) |  |
| Concomitant | 5973 (40.1%) | 5260 (71.0%) |  |
| **Outcomes** |  |  |  |
| Death | 445 (3.0%) | 616 (8.3%) | <0.001 |
| Hospitalized | 2936 (19.7%) | 2770 (37.4%) | <0.001 |
| Disability | 637 (4.3%) | 402 (5.4%) | <0.001 |
| **Source of report†** |  |  |  |
| Patient | 532 (3.5%) | 422 (5.7%) | <0.001 |
| Provider | 442 (3.0%) | 413 (5.6%) | <0.001 |

\*If > 1 reports for a case, used highest role (primary suspect > secondary suspect > interacting > concomitant)
†N=1457 (9.8%) of women, 1400 (18.8%) of men

Reporting rates, PRR, and ROR for statin-associated DM, and ROR reported in women vs. men are found in **Table 2** and summarized by sex and statin group in **Figure 2**. Compared with all other ADEs, statin-induced DM was disproportionately reported in all subjects (2.5 vs. 0.6%, PRR = 4.5), women only (2.9 vs. 0.6%, PRR = 5.2), and men only (1.5 vs. 0.6%, PRR = 2.4). This corresponded to a significant ROR of DM reporting for all statins of interest combined in women compared with men (ROR 1.9 [1.9-2.0], n=22,130 statin-associated DM ADEs).

**Table 2.** Signals of disproportionate reporting for statin-associated DM.

| Statin | Overall |  |  | Men |  |  | Women |  |  | Men vs. Women |
| --- | --- | --- | --- | --- | --- | --- | --- | --- | --- | --- |
|  | Reports | AE | PRR | Reports | AE | PRR | Reports | AE | PRR | ROR |
| All | 1,073,946 | 2.5% | 4.5* | 489,453 | 1.5% | 2.4* | 519,209 | 2.9% | 5.2* | 1.9 [1.9-2.0]<sup>†</sup> |
| Atorvastatin | 486,937 | 3.6% | 6.1* | 225,248 | 1.5% | 2.3* | 229,249 | 4.5 % | 7.6* | 3.0 [2.9-3.1]<sup>†</sup> |
| Simvastatin | 289,805 | 1.6% | 2.5* | 135,554 | 1.5% | 2.2* | 139,131 | 1.8% | 2.7* | 1.2 [1.2-1.3]<sup>†</sup> |
| Rosuvastatin | 188,567 | 2.4% | 3.7* | 83,659 | 2.0% | 2.9* | 92,986 | 2.4% | 3.5* | 1.2 [1.1-1.3]<sup>†</sup> |
| Pravastatin | 103,871 | 1.4% | 2.1* | 44,038 | 1.3% | 1.8 | 53,947 | 1.5% | 2.3* | 1.2 [1.1-1.4]<sup>†</sup> |

\*PRR significant for DM associated with statin (PRR > 2, $\chi^2 > 4$ , $\geq 3$ events)
<sup>†</sup>ROR significant in women vs. men significant (ROR [95% CI] > 1, $\geq 3$ events)

**Figure 2a.**
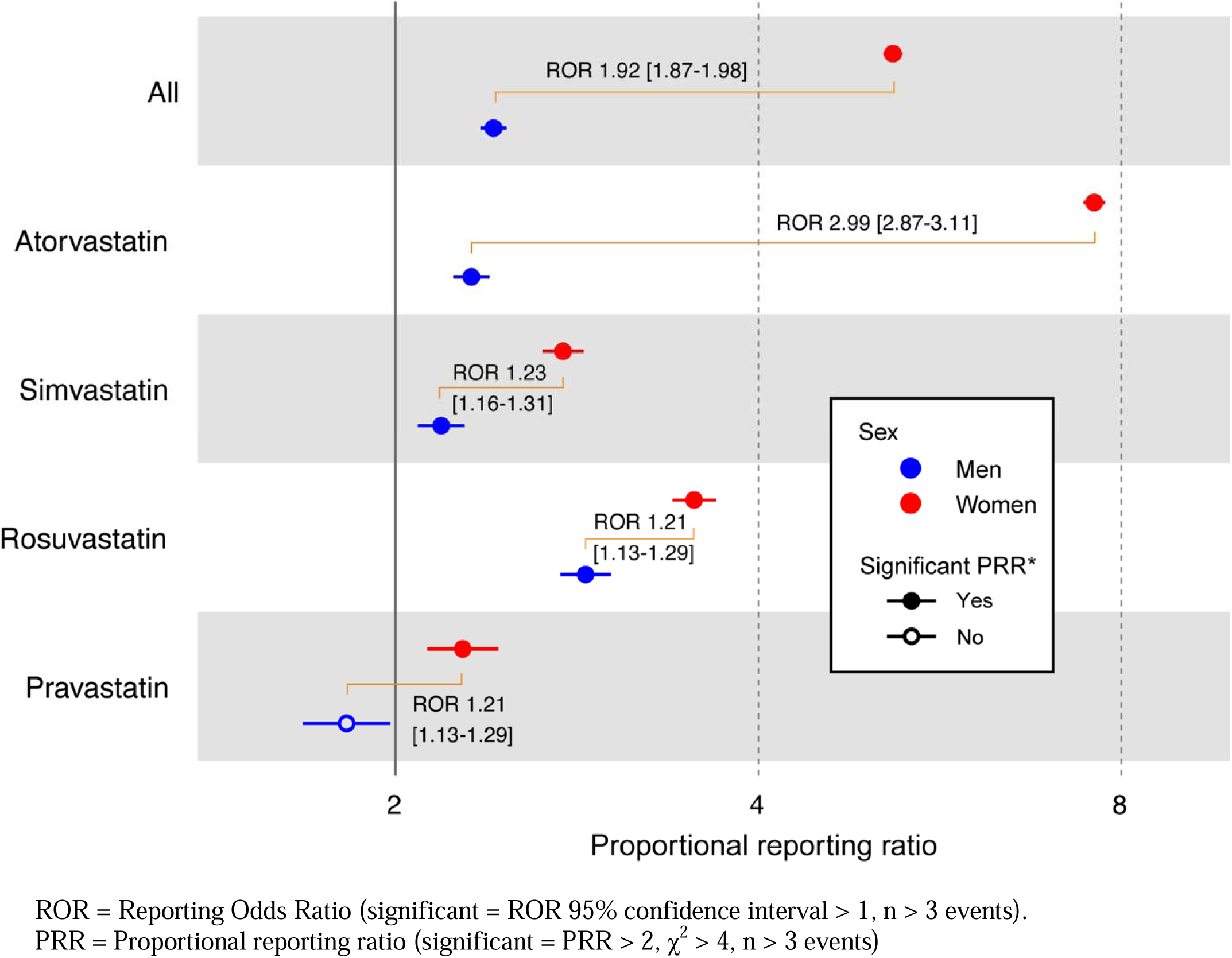
Sex-specific proportional reporting ratio for statin-associated DM.

**Figure 3b.**
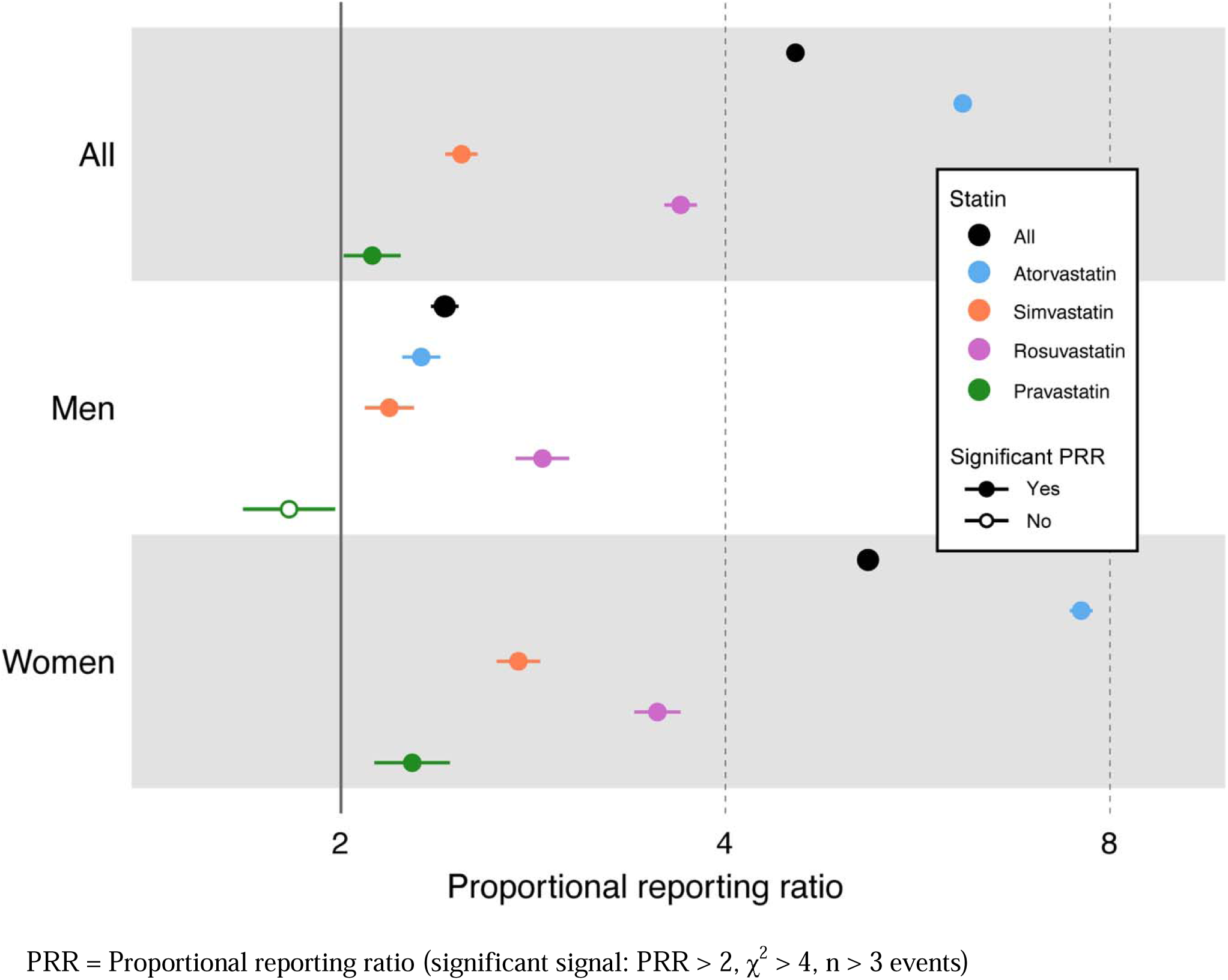
Statin-specific proportional reporting ratio for DM by sex.

DM was also reported with each individual statin (atorvastatin, simvastatin, rosuvastatin, pravastatin) at a higher rate compared with all other medications during the study period. Atorvastatin had the highest PRR (6.1), whereas pravastatin had the lowest (PRR 2.1). In men, the highest PRR for DM was associated with rosuvastatin (PRR 2.9), whereas in women the highest PRR was observed with atorvastatin (PRR 7.6). The PRR was numerically lowest for pravastatin in women (2.3) and was non-significant in men (1.8). The ROR indicated that the reporting rate in women was significantly larger for all statin groups compared to men with atorvastatin having the greatest difference (ROR 3.0 [95% CI 2.87-3.11]) and rosuvastatin having the smallest (ROR 1.2 [95% CI 1.13-1.29]).

## Discussion

### Principal findings

In 27 years of FAERS data, DM was reported at a significantly higher rate compared with all other drugs and at a significantly higher rate in women compared with men. Findings were similar for each of the 4 most reported individual statins, although the magnitude if difference between men and women was variable. Taken together, these data suggest a marked difference in the likelihood of statin-associated DM in women than in men with women.

### Comparison with other studies

The initial description from the Women’s Health Initiative of the differential risk of statin-associated DM between women and men was necessarily inferential given that only women were enrolled, and direct comparison between men and women in single datasets has proved challenging. Many large statin efficacy trials enrolled many more men than women, limiting statistical power to examine sex differences. Trials with a higher proportion of women such as the Stroke Prevention by Aggressive Reduction in Cholesterol Levels (SPARCL, atorvastatin, 40% women) study,^16^ the Justification for the Use of Statins in Prevention: an Intervention Trial Evaluating Rosuvastatin (JUPITER, 38% women),^17^ and Prospective Study of Pravastatin in the Elderly at Risk (PROSPER, 45% women)^18^ noted significantly increased risk of DM in the statin arms vs. placebo without stratification by sex, whereas studies enrolling predominately men did not. Male-predominant studies included the West of Scotland Coronary Prevention Study (WOCOPS, pravastatin, 0% women, OR of DM: 0.69 [0.49-0.96]),^19^ the Heart Protection Study (HPS, simvastatin, 25% women, OR of DM: 1.14 [0.98-1.33]),^20^ the Long Term Intervention with Pravastatin in Ischemic Disease (LIPID, pravastatin, 17% women, OR of DM: 0.95 [0.77-1.16]),^21^ Anglo-Scandinavian Cardiac Outcomes Trial (ASCOT, atorvastatin, 19% women, OR of DM: 1.14 [0.90-1.43]),^22^ and the Controlled Rosuvastatin Multinational Study in Heart Failure (CORONA, rosuvastatin, 24% women, OR of DM: 1.13 [0.86-1.49]).^23^ Our post-marketing data taken together with these clinical trial data are consistent with a higher risk of DM due to statins in women than men.

We observed that the percentage of statin-related ADEs involving women vs. men was similar (52% women). Since women are less likely to be prescribed statin therapy than men even when indicated,^5,7,28^ it is likely that the relative reporting rate of statin-associated ADE per patient is even higher in women than the number of ADE reports would suggest. Prior studies have demonstrated higher rates of ADE reporting in women compared with men.^29,30^ This has been observed across multiple drug classes and regions including the European Union’s Vigibase,^31^ Sweden,^32^ Africa,^33^ the Netherlands,^34^ FAERS,^15,35^ and in meta-analyses of published clinical trials.^36^ Interestingly, although ADEs are reported more frequently in women than men, the severity of ADE complications with respect to outcomes like hospitalization, disability, and death are often higher in men.^31,37^ It has been speculated that sex differences in metabolism and pharmacokinetics may contribute to the higher rates of ADEs in women vs. men we observed in our analysis, suggesting that targeted studies and possibly specific dosing recommendations should be considered.^38^ Other factors besides pharmacokinetics may also contribute to sex differences in ADE frequency, such as gene and protein expression of drug targets or differences in the likelihood of patients and providers reporting a given ADE. We attempted to mitigate potential reporting bias by directly comparing women vs. men using the modified ROR, which compares the frequencies of a specific ADE (e.g., DM) with all ADEs for each subgroup. By this method, women still demonstrated a higher risk of DM than men with all statins, particularly atorvastatin.

### Strengths and limitations

To our knowledge, this is the first analysis of real-world, post-marketing surveillance data to investigate and demonstrate sex differences in statin-associated DM. Observational studies and pooled analyses of randomized clinical trials have suggested the risk of statin-associated DM is ‘intensity-dependent’ (simvastatin, atorvastatin, rosuvastatin over pravastatin and fluvastatin), although sex differences have not been previously evaluated.^24–26^ We found that atorvastatin appeared to have the greatest SDR for DM overall (PRR 6.1) and the largest difference between women and men (ROR 3.0 [95% CI 2.9-3.1]). The next largest overall SDRs were associated with rosuvastatin (PRR 3.7 overall, women vs. men ROR 1.2 [95% CI 1.1-1.3]) and simvastatin (PRR 2.5 overall, women vs. men ROR 1.2 [95% CI 1.2-1.3]). These findings are concordant with prior meta-analyses in which all three agents appeared associated with DM.^27^ Pravastatin was associated with the smallest SDR overall (PRR 2.1) in our analysis though still with a significant difference between women and men similar to rosuvastatin and simvastatin (ROR 1.2 [95% CI 1.1-1.4]). This last finding is concordant with previous secondary analyses of randomized clinical trials, which have noted a neutral or even reduced risk of DM with pravastatin.^27^ The relative significance of SDRs among the statins in our analysis is concordant with prior prospective studies in terms of overall trends. Numeric differences between PRR in women and men in our study were also larger in atorvastatin, rosuvastatin, and simvastatin compared with pravastatin as has been observed previously. Our results also suggest that women may have a higher propensity for statin-associated DM than men, although the magnitude of difference may vary among statin agents.

Our study had several limitations common to pharmacovigilance analyses. FAERS is a spontaneous reporting database wherein only a small fraction of all ADEs that occur are actually captured. Consequently, point estimates of actual ADE frequency cannot be derived using spontaneous reporting systems like FAERS, and observed differences in ADE reporting rates between men and women may be affected by numerous confounding factors.^39^ Sources of ADE reports include patients, providers, payors, and pharmaceutical manufacturers and can be impacted by proximity to FDA approval, publication of possible ADE associations, and interactions between clinicians and patients. Bias, including sex differences, can occur regarding which ADEs are submitted to FAERS.^37^ FAERS reports are generally not adjudicated, and classification of a specific adverse effect (e.g. DM) may be inaccurate. Finally, SDRs reflect only relative reporting of ADEs between different populations and cannot be taken as direct evidence of an increase in absolute rate of an ADR. Therefore, these findings should be taken as a) supportive evidence of prior suspected associations and b) foundational data on which to design future translational or observational studies.

## CONCLUSIONS

Using 27 years of real-world post-marketing spontaneous ADE reporting data, we found that disproportionate reporting of statin-associated DM was significantly higher in women than in men. Significant sex differences were observed with every statin analyzed but were greatest with atorvastatin. Future studies should elucidate the mechanism of these differences to determine which statins might be safer in treating women compared to men.

## Data availability statement

The raw data used in this study is publicly available at https://open.fda.gov/data/faers/). The drug names and ingredients of FDA approved products is available at https://www.fda.gov/drugs/drug-approvals-and-databases/drugsfda-data-files. The MedDRA terminology is available with a license at https://www.meddra.org/. The R mdsstat analysis package is at https://rdrr.io/cran/mdsstat/. Analysis code and access to our FAERS BigQuery repository containing linked versions of the above resources are available on reasonable request.

